# A systematic review and meta-analysis considering the SSRI, SNRI and TCA classes of antidepressants and the risk for congenital heart defects

**DOI:** 10.1101/2020.02.24.20027532

**Authors:** Courtney De Vries, Svetla Gadzhanova, Matthew J. Sykes, Michael Ward, Elizabeth Roughead

## Abstract

**Background:** Antidepressant use during the first trimester is reported in 4% to 8% of pregnancies. The use of some selective serotonin reuptake inhibitors (SSRI) during this stage of gestation has been identified as increasing the odds for congenital heart defects, however little is known about the safety of non-SSRI antidepressants.

**Objective:** To assess the odds of congenital heart defects associated with the use of any antidepressant during the first trimester of pregnancy. To investigate individual classes of antidepressants: SSRIs, serotonin norepinephrine reuptake inhibitors (SNRI), tricyclic antidepressants (TCA) and individual antidepressants.

**Data sources:** PubMed and Embase were searched without restrictions from inception till 2 January 2020.

**Study selection:** Prospective and retrospective cohort and case-control studies were included if they documented the maternal usage of antidepressants during the first trimester of pregnancy and assessed the presence of congenital heart defects.

**Data extraction and meta-analysis:** Data were extracted by two independent reviewers and the endpoint assessed was congenital heart defects. Where studies reported multiple results for different types of heart defects or individual antidepressants, results were combined when possible. Analyses assessing individual antidepressants and classes of antidepressants (SSRIs, SNRIs and TCAs) were undertaken.

**Results:** A total of 16 studies were identified, encompassing 4,564,798 pregnancy outcomes. The odds ratio for maternal use of any antidepressant and the presence of congenital heart defects from the mixed-methods meta-analysis was 1.22 (95% confidence interval (CI): 1.11 to 1.33).

Analyses of antidepressants by class produced an odds ratio of 1.50 (95% CI: 1.19 to 1.89) for maternal SNRI use during the first trimester of pregnancy and the formation of congenital heart defects. A significant odds ratio of 1.22 (95% CI: 1.12 to 1.33) was reported for SSRIs. For the TCA class, no increased odds ratio was found.

Analyses of individual antidepressants produced significant odds ratios of 1.53 (95% CI: 1.25 to 1.88), 1.28 (95% CI: 1.01 to 1.62), 1.28 (95% CI: 1.14 to 1.45) and 1.23 (95% CI: 1.01 to 1.50) for paroxetine, fluoxetine, sertraline and bupropion respectively.

**Conclusion:** While some insight has been gained into which classes of antidepressant and individual antidepressants pose more risk than others for causing congenital heart defects, information regarding some antidepressants is still lacking.

## Introduction

Antidepressants are used during pregnancy, with the prevalence of maternal antidepressant use increasing over time1^1-3^. In Denmark, the use of selective serotonin reuptake inhibitors (SSRI) in pregnancy increased from approximately 0.5% to 3% between 2000 and 2010.^1^ In the United States of America (USA), the Birth Defects Study identified that the use of any antidepressant during the first trimester of pregnancy increased from less than 0.5% before 1987, to 3% in 1999 and 8% in 2008.^2^ This study identified that use of selective serotonin reuptake inhibitors (SSRI) was 2% in 1999 and 6% in 2008.^2^ Dispensing records between 2006 and 2011 in the USA, identified the use of any antidepressant and SSRIs during the first trimester of pregnancy as 5% and 4% respectively.^3^

Paroxetine was the first SSRI to reach the American market in 1992. In 2005, an alert was released stating that the use of paroxetine during pregnancy had been associated with an increased risk of major congenital malformations, especially those relating to the heart^4^. A 2016 meta-analysis reported an odds ratio of 1.23 (95% confidence interval (CI): 1.06 to 1.43) for paroxetine use during the first trimester of pregnancy and any cardiac malformations.^5^ Specific cardiac defects were also investigated in this meta-analysis and some were reported to be significant, including: atrial septal defects, right ventricular outflow tract obstructions, bulbus cordis anomalies and anomalies of cardiac septal closure.^5^

Medications from the same class may exert similar toxicities, meaning that other antidepressants may also increase the risk for congenital heart defects. It is desirable to minimise medicine use in pregnancy but discontinuing antidepressant therapy in unwell women may be complicated by the risks of suicidal ideation, relapse of depression and anxiety or panic attacks.^6 7^ The severity of these risks mean that it may not be possible to cease antidepressant therapy. Evidence regarding the safety of antidepressant use in human pregnancies is required for all women.

Pharmacoepidemiological studies provide data for birth outcomes following antidepressant use during the first trimester of pregnancy, which is the relevant period to investigate for cardiac defects as the heart is formed during weeks 4 to 8 of gestation. Previous meta-analyses have revealed significant relationships between usage of paroxetine,^5 8 9^ fluoxetine,^10 11^ sertraline^12^ and any antidepressant^13^ during the first trimester of pregnancy and an increased formation of heart defects. Only one of these meta-analyses considered non-SSRI antidepressants,^13^ and several cohort and case-control studies have been published since the last published meta-analysis.^14-18^ The largest meta-analysis to date was carried out by Shen et al.^12^ and investigated the first trimester use of sertraline and congenital heart defects. This meta-analysis included 12 cohort studies with a total of 6,468,241 pregnant women and found a statistically significant increased risk of cardiovascular defects, with an odds ratio of 1.36 (95% CI: 1.06 to 1.74).

The aim of this systematic literature review and meta-analysis was to investigate the relationship between usage of any antidepressant during the first trimester of pregnancy and the development of congenital heart defects. An investigation into specific classes of antidepressants was also completed to assess whether a class-effect exists between first trimester antidepressant use and congenital heart defects. The relationship between usage of specific antidepressants during the first trimester of pregnancy and heart defects was also investigated.

## Methods

### Literature review

A literature review was completed using PubMed and Embase. The databases were searched from inception to 2 January 2020 using MeSH terms listed in supplement 1. Search terms were searched independently and then combined within each database to retrieve articles reporting the use of antidepressive medications in pregnancy and adverse outcomes. The search strategy and results are detailed in supplement 1. The results were limited to human data and no further limits were imposed on the search. Duplicates were removed before moving onto the title and abstract reviews. PRISMA guidelines were followed for all procedures and reporting (supplement 2).

### Study selection criteria and data extraction

The title and abstract inclusion criteria were cohort or case-control studies with a comparison group of women who had not used antidepressants during pregnancy, study participants aged 15 years or older, maternal use of antidepressants during the first trimester of pregnancy, and outcomes which included congenital heart defects or other malformations. Live births, stillbirths and terminations of pregnancy were all classified as suitable endpoints of pregnancies. If the usage of antidepressant agents was not confirmed, repeated prescriptions or dispensations of an agent were assumed to correlate directly to maternal usage.

The title and abstract exclusion criteria were: reviews, study protocols, case series, case reports, editorials, commentaries, letters to the editor, conference papers, study participants under 15 years of age, studies which contained only abortive, mortal and neurobehavioral outcomes and if the data collected did not pertain to antidepressant medications. The form for title and abstract reviews is provided in supplement 3.

The quality of eligible studies was assessed using a modified version of the Newcastle-Ottawa quality assessment scale. While the Newcastle Ottawa Scale assesses important study features it is not intuitive to use and reviews have documented that there is a low inter-reviewer reliability and poor agreement about study quality between authors and reviewers when using this scale.^19 20^ For this reason the scale was altered to make it tailored for assessing congenital heart defects as the outcome of interest and to improve categorisation of study design. To improve the ease of which reviewers could assess study design the scale was converted to a tabular format which allowed for an analysis on the study quality to be compiled into a single document. The advantage of this method is that it allowed customisation of the documentation of the study methods before grouping the criteria into high, medium or low categories and the assessment outcomes into ideal, good, acceptable or poor outcomes (supplement 4). High impact criteria assessed the selection of study participants and the determination of outcomes and antidepressant use. Follow-up was classified as being a medium impact criterion. Low impact criteria assessed other study features including maternal smoking, alcohol or medication use and whether the non-medicated comparison group was depressed or not.

If three of the high impact criteria were ideal, the end of follow-up was not poor and at least one of the low impact criteria were ideal for cohort studies, the study was included. Cohort studies were excluded if any of the high or medium impact criteria were poor. If four of the high impact criteria were ideal and at least two of the low impact criteria were ideal for case-control studies, the study was included. Case-control studies were excluded if any of the high impact criteria were poor. The differences in cohort and case-control study designs produced slightly different study criteria and therefore inclusion decisions. The method for ascertaining antidepressant use, a high impact criterion, was completed for both cohort and case-control studies. For case-control studies an additional criterion was added to confirm that the same method of ascertainment was used for cases and controls. End of follow-up, a medium impact criterion, was only completed for cohort studies to assess pregnancy and birth outcomes. Non-response rate, a low impact criterion, was only completed for case-control studies to identify any differences between the non-response rates of cases and controls (supplement 4).

CDV and SG independently undertook data extraction and quality ranking. Any conflicts were discussed and reviewed by ER.

Once potentially suitable studies were identified, the data sources, study years and settings were compared to ensure that multiple studies did not use the same data. If data sources such as birth registries overlapped between studies, then the most recent study providing the largest sample size was retained.

All reported crude odds ratios and adjusted odds ratios were extracted from the studies. Additionally, the data needed to calculate odds ratios for antidepressant usage during the first trimester of pregnancy and the formation of congenital heart defects were retrieved from the studies.

Most studies included a single comparison group which had not used antidepressants during pregnancy, however some studies reported multiple comparison groups. Ban, et al. 2014^14^ provided data on two unmedicated groups, either depressed or non-depressed. The comparison group in the meta-analysis comprises the sum of unmedicated depressed and non-depressed women. Furu, et al 2015^16^ provided data on a full study cohort and a sub-cohort comprised of sibling pairs. Data from the full cohort were included. Davis et al. 2007^21^ reported data on SSRIs and tricyclic antidepressants (TCA), however these could not be combined for the primary analysis because the total number of women in the unmedicated comparison group was inconsistent. Therefore, these data are reported individually in all analyses.

Some studies assessed individual heart defects or individual antidepressants. Whenever possible, results from the individual antidepressants and heart defects were aggregated to estimate overall harm for the “any antidepressant” analysis. Classes of antidepressant and individual antidepressants were analysed if data for the meta-analyses were available from at least three studies for each class or antidepressant.

Confounding factors reported in the studies including the use of other medications, maternal smoking and maternal diabetes were recorded.

### Statistical analyses

RevMan 5.3^22^ was utilised to perform the meta-analyses. The primary analysis investigated the relationship between maternal use of any antidepressant during the first trimester of pregnancy and the presence of congenital heart defects. The antidepressant classes were stratified into SSRI, serotonin-norepinephrine reuptake inhibitor (SNRI) and TCA groups. Antidepressants were investigated individually, and the results were also stratified by cohort and case-control studies.

Mixed-effects models were used for all meta-analyses because they are more suited to dealing with heterogenous data. An overall odds ratio with a 95% CI measuring the relationship between first trimester antidepressant usage and the presence of heart defects was calculated in all analyses and the significance of these relationships were investigated by the p value for overall effect. Heterogeneity was measured by the I^2^ values and their corresponding p values and analyses were defined as being heterogenous if their p values were significant (<0.05).

Outlying studies were defined as having a markedly different intervention effect estimate or an abnormally large confidence interval. To complete a sensitivity analysis, the primary meta-analysis was investigated for outliers and a funnel plot was examined to identify outlying studies (supplement 5). The funnel plot is also a small-study effect test. The Mantel-Haenszel mixed-effects meta-analysis was applied, both with and without the outlying studies, to determine the effect of outliers on the overall significance of the results and the heterogeneity.

## Results

Figure 1 shows the article selection process. Following the title and abstract screening, 76 articles were eligible for a full text review. 16 studies, comprising 4,564,798 pregnancy outcomes met the final inclusion criteria and were included.

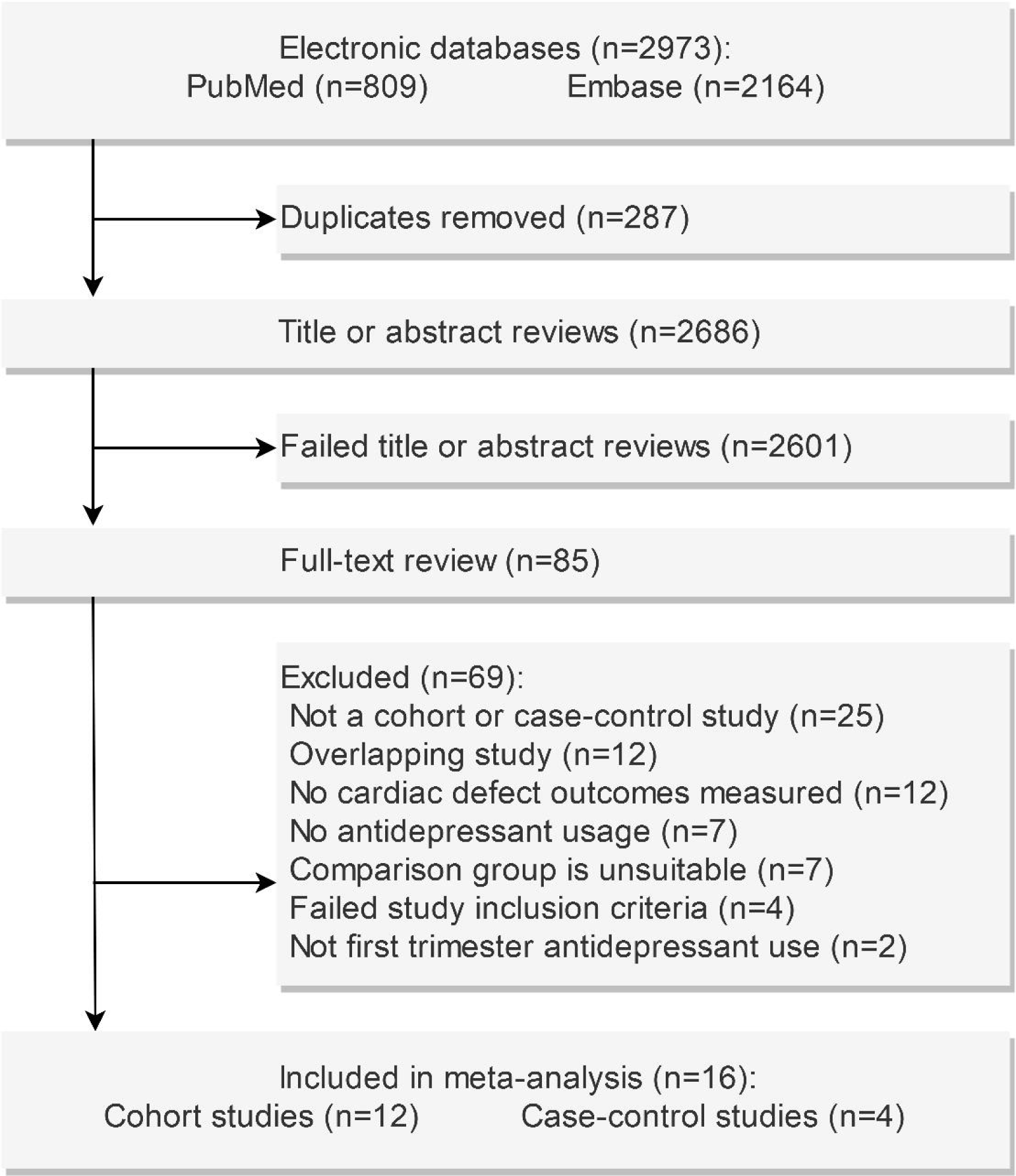

An overview of the characteristics of the included studies and detailed information on relevant medication use is reported in table 1. Twelve studies were cohort studies ^5 14 16 21 23-29^and four studies were case controls. ^18 30-32^ Eight of the studies were prospective^14-16 18 24 27 28 33^ and eight were retrospective. ^21 23 25 26 29-32^ For two studies the comparison group included depressed women who were unmedicated. ^14 15^

**Table 1.** Overview of studies included in the systematic review and meta-analyses

| **Author, Year Published** | **Study Type** | **Study Years (Setting)** | **End Of Follow Up** | **Definition Of Maternal Antidepressant Use** | **Definition Of Non-Antidepressant Use** | **Proportion Of Cohort With Concomitant Medications** |
| --- | --- | --- | --- | --- | --- | --- |
| Jordan, et al. 2016<sup>24</sup> | Prospective cohort | 2004 – 2010 (Norway), 2000 – 2010 (Wales and Denmark) | Live births, still births, fetal deaths after 20 weeks (24 weeks in Wales) | One or more prescription for an antidepressant (Wales) or dispensed (Norway and Denmark), 91 days either side of the first day of the LMP | Did not use any antidepressant during the 91 days either side of the first day of the LMP | n.d. |
| Furu, et al. 2015<sup>16</sup> | Prospective cohort | 1996 – 2010 (Denmark, Finland, Iceland, Norway and Sweden) | Live births only | Use of a single, consistent SSRI between at least one month before conception and day 84 of the pregnancy, identified by redeemed prescriptions | No redeemed prescriptions for antidepressants from 30 days before the first day of the LMP to 97 days after the LMP | 15% of SSRI-using and 1% of non-SSRI-using mothers |
| Berard, et al. 2015<sup>15</sup> | Prospective cohort | 1998 – 2010 (Canada) | Live births only | Continuous prescription of antidepressants for at least 12 months before the LMP and during pregnancy | Depressed and not using antidepressant medications | 93% of mothers using sertraline<br>93% of mothers using non-sertraline SSRIs<br>94% of mothers using non-SSRI antidepressants<br>84% of mothers in the comparison group |
| Huybrechts, et al. 2014<sup>23</sup> | Retrospective cohort | 2000 – 2007 (United States) | Live births only | Overlap between pharmacy dispensing of antidepressants and the first trimester of pregnancy | No dispensing of antidepressants during the first trimester of pregnancy | Among SSRI users: 4% used antidiabetics, 16% used anticonvulsants, 21% used antipsychotics, 7% used anxiolytics, 32% used benzodiazepines, 29% used other hypnotic agents, 8% used barbiturates and 8% used a suspected teratogen |
| Ban, et al. 2014<sup>14</sup> | Prospective cohort | 1990 – 2009 (United Kingdom) | Live births only | Presence of SSRI or TCA prescriptions in records from four weeks before to 12 weeks after the first day of the estimated LMP. Dual usage of SSRIs excluded | Depressed or non-depressed and not using antidepressants | n.d. |
| Vasilakis-Scaramozza, et al. 2017<sup>29</sup> | Retrospective cohort | 1991 – 2002 (United Kingdom) | Live births, stillbirths, and therapeutic abortions > 20 weeks' gestation | Prescription receipt for an antidepressant from 180 to 335 days prior to the delivery date for live births and from 70 to 225 days prior to the delivery date for stillbirths and therapeutic abortions | No receipt for an antidepressant during pregnancy | Less than 1% of women using an antidepressant used insulin or a teratogen during the first trimester of pregnancy |
| Margulis, et al. 2013<sup>26</sup> | Retrospective cohort | 1996 – 2010 (United Kingdom) | Live births only | Women with one or more prescriptions for a single SSRI, overlapping with the first trimester | Women with no antidepressant prescriptions in the three | Among SSRI users: 18% had any prescription for non-SSRI psychotropics, 1% had any |

|  |  |  |  | of pregnancy | months before through to the first or second trimester of pregnancy | prescriptions for anticonvulsants in and 4% had non-antidepressant drugs prescribed in the baseline year |
| --- | --- | --- | --- | --- | --- | --- |
| Nordeng, et al. 2012<sup>27</sup> | Prospective cohort | 1999 – 2005 (Norway) | Live births, fetal deaths, or induced abortions after gestational week 12 | Dispensed prescription for any antidepressant from 30 days before the first day of the LMP until 97 days after the LMP | No reported use of any antidepressants in the six months before or during pregnancy | n.d. |
| Diav-Citrin, et al. 2008<sup>33</sup> | Prospective cohort | 1994 – 2002 (Israel and Italy), 2002 – 2005 (Germany) | Live births only | Dispensed prescription for paroxetine or fluoxetine from 30 days before the first day of the LMP till 97 days after the LMP | Women who contacted a centre regarding the use of medications known not to be harmful in similar timeframes | n.d. |
| Oberlander, et al. 2008<sup>28</sup> | Prospective cohort | 1998 – 2001 (Canada) | Live births only | Dispensed antidepressant prescription from the LMP to 90 days after the LMP | No dispensed prescriptions for antidepressants or benzodiazepines | Women taking anticonvulsants were excluded |
| Davis, et al. 2007<sup>21</sup> | Retrospective cohort | 1996 – 2000 (United States) | Live births only | Dispensed antidepressant during the first trimester of pregnancy | Women who were not prescribed antidepressants at any time during pregnancy | n.d. |
| Kulin, et al. 1998<sup>25</sup> | Retrospective cohort | Unknown time period (United States and Canada) | Live births, spontaneous abortions, therapeutic abortions and stillbirths | Use of sertraline or fluoxetine during the first trimester of pregnancy | Use of non-harmful medications during pregnancy | n.d. |
| Louik, et al. 2014<sup>18</sup> | Prospective case-control | 1993 – 2004 (United States) | Live births only | Use of any antidepressant from 28 days before the LMP to 112 days after the LMP | No use of any antidepressant at any time from 56 days prior to the LMP | n.d. |
| De Jonge, et al. 2013<sup>31</sup> | Retrospective case-control | 1998 – 2008 (Netherlands) | Live births, stillbirths, spontaneous abortions and terminations of pregnancy | The antidepressant was registered to be used during the first 13 weeks after the first day of the LMP | The antidepressant was not registered to be used during the first 13 weeks after the first day of the LMP | n.d. |
| Polen, et al. 2013<sup>32</sup> | Retrospective case-control | 1997 – 2007 (United States) | Live births, still births ≥ 20 weeks gestation and elective terminations | Reported use of venlafaxine from one month before conception through to the third month of pregnancy | No reported use of any antidepressant from three months before conception through the end of pregnancy | n.d. |
| Alwan, et al. 2010<sup>30</sup> | Retrospective case-control | 1997 – 2004 (United States) | Live births and fetal deaths after 20 weeks gestation | Reported use of bupropion anytime between one month before and three months after conception | Did not use any antidepressant at any time during pregnancy | n.d. |

LMP, Last menstrual period; n.d., no data; SSRI, Selective serotonin reuptake inhibitor; TCA, tricyclic antidepressant

The unadjusted and adjusted odds ratios from individual studies are provided in table 2, with significant odds ratios shown in bold text.

**Table 2.** Systematic review of the individual studies, reporting antidepressant use during the first trimester of pregnancy and odds ratios for congenital heart defects

| Author, Year Published | Cohort / Case |  | Comparison |  | Antidepressant Used | Crude Odds Ratio (95% CI) | Adjusted Odds Ratio (95% CI) | Odds Ratio Adjusted For | Meta Analysis That The Data Was Used In |
| --- | --- | --- | --- | --- | --- | --- | --- | --- | --- |
|  | N Events | N Total | N Events | N Total |  |  |  |  |  |
| Jordan, et al. 2016<sup>24</sup> | 121 | 12,962 | 4,503 | 506,155 | SSRI or SNRI | 1.03 (0.86 to 1.24) | 1.00 (0.82 to 1.21) | Smoking and socioeconomic status | Any antidepressant |
|  | 101 | 10,959 | 4,047 | 449,643 | SSRI monotherapy | 1.00 (0.82 to 1.22) | n.d. | n.d. | SSRI group |
|  | 18 | 1,448 | 4,503 | 506,155 | SNRI | 1.40 (0.88 to 2.23)* |  |  | SNRI group |
| Furu, et al. 2015<sup>16</sup> | 530 | 34,535 | 26,745 | 2,266,875 | SSRI or venlafaxine | **1.32 (1.21 to 1.44)\*** | n.d. | n.d. | Any antidepressant |
|  | 490 | 31,772 |  |  | SSRI | **1.33 (1.21 to 1.45)\*** |  |  | SSRI group |
|  | 40 | 2,763 |  |  | Venlafaxine (monotherapy or polytherapy) | 1.23 (0.90 to 1.68) | 1.14 (0.82 to 1.57) | Maternal age, year of birth, birth order, smoking, maternal diabetes, country and use of other medications | SNRI group |
| Berard, et al. 2015<sup>15</sup> | 89 | 3,625 | 344 | 14,868 | Any antidepressant | 1.06 (0.84 to 1.35)* | n.d. | n.d. | Any antidepressant |
|  | 61 | 2,329 |  |  | SSRI | 1.14 (0.86 to 1.50)* |  |  | SSRI group |
| Huybrechts, et al. 2014<sup>23</sup> | 580 | 64,389 | 6,403 | 885,115 | Any antidepressant | **1.25 (1.15 to 1.36)** | n.d. | n.d. | Any antidepressant |
|  | 416 | 46,144 |  |  | SSRI | **1.25 (1.13 to 1.38)** |  |  | SSRI group |
|  | 75 | 6,904 |  |  | SNRI | **1.51 (1.20 to 1.89)** |  |  | SNRI group |
|  | 42 | 5,954 |  |  | TCA | 0.97 (0.72 to 1.32) |  |  | TCA group |
| Ban, et al. 2014<sup>14</sup> | 90 | 10,401 | 2,556 | 338,726 | SSRI or TCA mono- or polytherapy | 1.15 (0.93 to 1.42)* | n.d. | n.d. | Any antidepressant |
|  | 68 | 7,683 |  |  | SSRI monotherapy | 1.17 (0.92 to 1.50)* |  |  | SSRI group |
|  | 20 | 2,428 |  |  | TCA monotherapy | 1.09 (0.70 to 1.70)* |  |  | TCA group |
| Vasilakis-Scaramozza, et al. 2013<sup>29</sup> | 18 | 3,287 | 28 | 6,617 | SSRI or TCA | 1.30 (0.72 to 2.35)* | n.d. | n.d. | Any antidepressant |
|  | 7 | 1,825 |  |  | SSRI | 0.91 (0.40 to 2.08)* |  |  | SSRI group |
|  | 11 | 1,608 |  |  | TCA | 1.62 (0.81 to 3.26)* |  |  | TCA group |
| Margulis, et al. 2013<sup>26</sup> | 16 | 3,046 | 48 | 8,991 | SSRI | 0.98 (0.56 to 1.73)* | 1.09 (0.66 to 1.79) | Logistic regression | Any antidepressant and SSRI group |
| Nordeng, et al. 2012<sup>27</sup> | 6 | 556 | 541 | 61,648 | Any antidepressant | 1.23 (0.55 to 2.77)* | 1.24 (0.55 to 2.82) | Maternal depression, maternal age at delivery, parity and use of psychotropic medications during pregnancy | Any antidepressant |
|  | 6 | 462 |  |  | SSRI | 1.49 (0.66 to 3.34)* | 1.51 (0.67 to 3.43) |  | SSRI group |
| Diav-Citrin, et al. 2008<sup>33</sup> | 14 | 601 | 8 | 1,359 | Paroxetine or fluoxetine | **4.03 (1.68 to 9.65)\*** | n.d. | n.d. | Any antidepressant and SSRI group |
| Oberlander, et al. 2008<sup>28</sup> | 19 | 2,709 | 512 | 107,320 | SSRI or SNRI monotherapy | 1.47 (0.93 to 2.33)* | n.d. | n.d. | Any antidepressant |
|  | 18 | 2,459 |  |  | SSRI monotherapy | 1.54 (0.96 to 2.47)* |  |  | SSRI group |
|  | 1 | 250 |  |  | Venlafaxine monotherapy | 0.84 (0.12 to 5.98)* |  |  | SNRI group |
| Davis, et al. 2007<sup>21</sup> | 17 | 805 | 1,171 | 49,031 | SSRI | 0.88 (0.54 to 1.43)* | n.d. | n.d. | Any antidepressant and SSRI group |
|  | 2 | 167 | 1,186 | 49,669 | TCA | 0.50 (0.12 to 2.00)* |  |  | Any antidepressant and TCA group |
| Kulin, et al. 1998<sup>25</sup> | 2 | 267 | 4 | 267 | SSRI | 0.50 (0.09 to 2.73)* | n.d. | n.d. | Any antidepressant and SSRI group |
| Louik, et al. 2014<sup>18</sup> | 447 | 8,805 | 370 | 8,611 | Any antidepressant | **1.19 (1.03 to 1.37)\*** | n.d. | n.d. | Any antidepressant |
|  | 349 |  | 290 |  | SSRI | **1.18 (1.01 to 1.39)\*** |  |  | SSRI group |
|  | 29 |  | 32 |  | TCA | 0.89 (0.54 to 1.47)* |  |  | TCA group |
| De Jonge, et al. 2013<sup>31</sup> | 14 | 873 | 281 | 29,223 | SSRI | 1.68 (0.98 to 2.88)* | n.d. | n.d. | Any antidepressant and SSRI group |
| Polen, et al. 2013<sup>32</sup> | 74 | 15,758 | 14 | 8,002 | Venlafaxine | **2.69 (1.52 to 4.77)\*** | n.d. | n.d. | Any antidepressant and SNRI group |
| Alwan, et al. 2010<sup>30</sup> | 34 | 6,853 | 26 | 5,869 | Bupropion | 1.12 (0.67 to 1.87)* | 1.4 (0.8 to 2.5) | Maternal race, obesity, smoking and family income | Any antidepressant |

CI, confidence interval; SSRI, selective serotonin reuptake inhibitor; SNRI, serotonin norepinephrine reuptake inhibitor; n.d., no data; TCA, tricyclic antidepressant
\*Odds ratio and 95% CI calculated using RevMan 5.3

The total pooled odds ratio from the unadjusted meta-analysis for any antidepressant use was 1.22 (95% CI: 1.11 to 1.33, p < 0.0001) (figure 2). The corresponding number needed to harm is 442 women taking antidepressants during the first trimester of pregnancy.

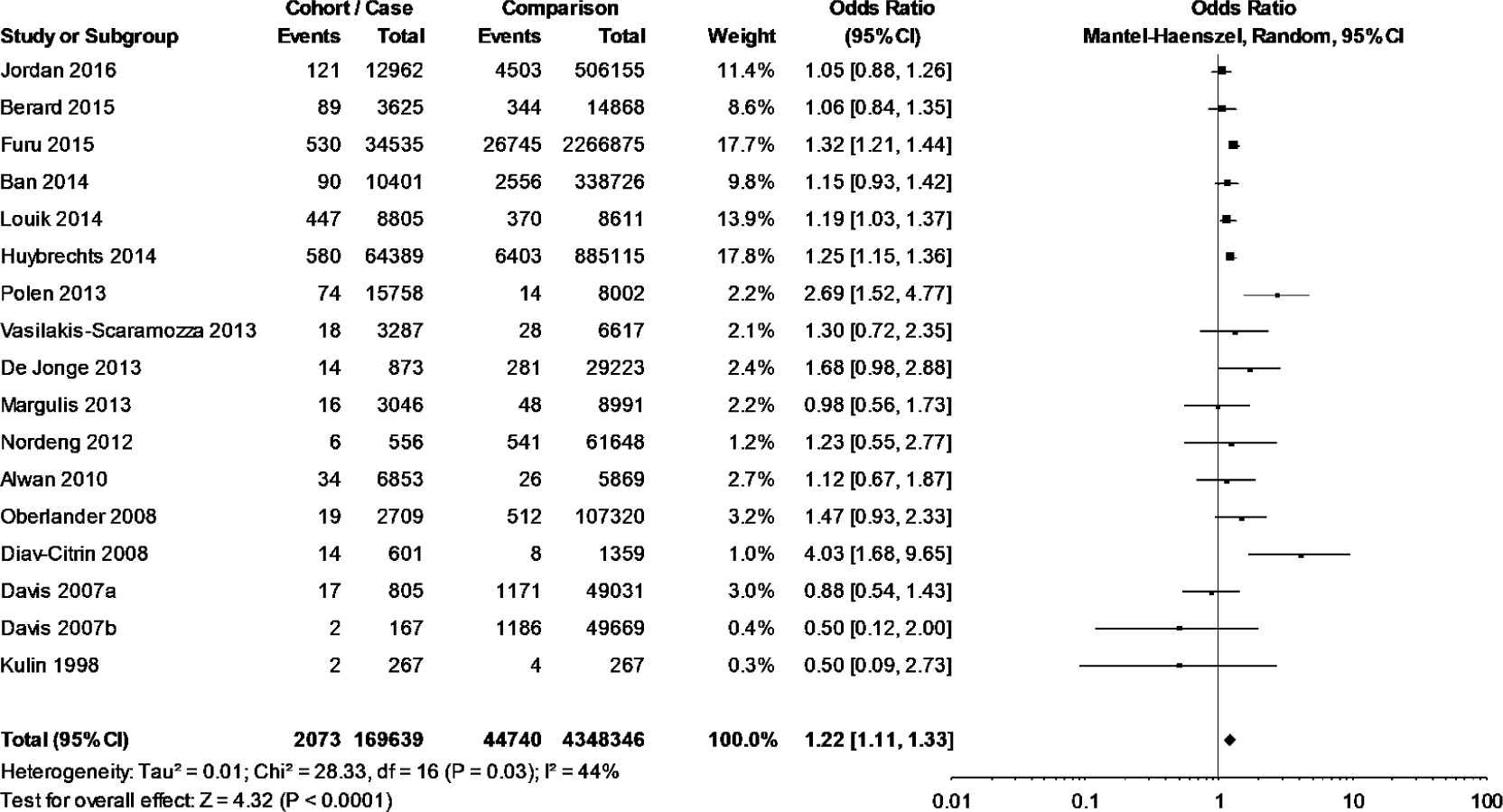

Statistically significant odds ratios of 1.22 (95% CI: 1.12 to 1.33, p < 0.00001) and 1.50 (95% CI: 1.19 to 1.89, p < 0.0006) are reported in figure 3 for the SSRI and SNRI subgroups respectively. The corresponding numbers needed to harm are 198 and 447 women taking SSRIs or SNRIs during the first trimester of pregnancy. The TCA subgroup had a non-statistically significant odds ratio of 1.01 (95% CI: 0.82 to 1.25, p = 0.52) (figure 3). Exclusion of the outliers did not change the significance of the results for the overall or class analyses (supplement 5).

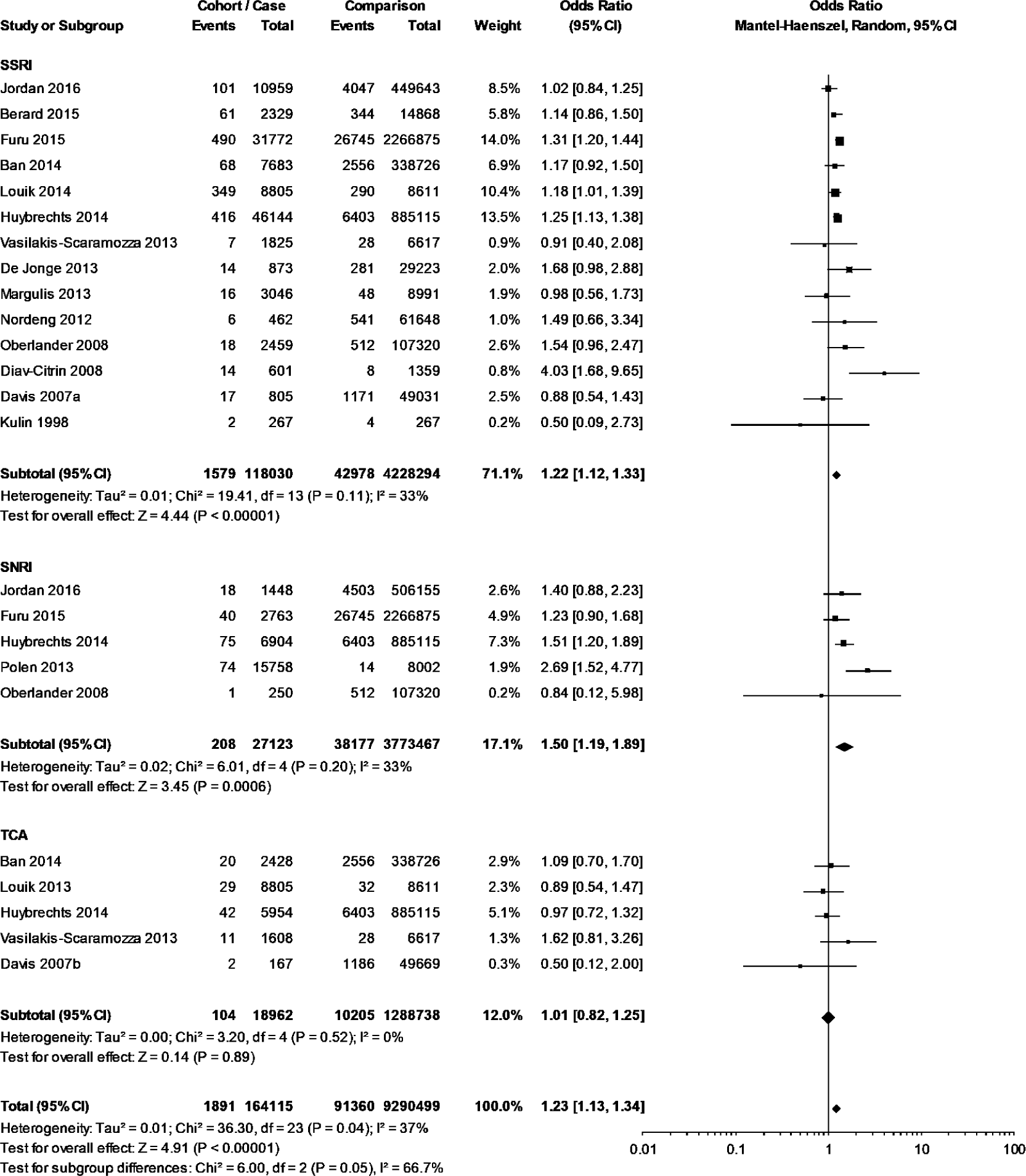

Analyses of individual antidepressants produced statistically significant odds ratios of 1.53 (95% CI: 1.25 to 1.88, p < 0.001) for paroxetine, 1.28 (95% CI: 1.01 to 1.62, p = 0.04) for fluoxetine, 1.28 (95% CI: 1.14 to 1.45, p < 0.0001) for sertraline and 1.23 (95% CI: 1.01 to 1.50, p = 0.04) for bupropion (figure 4). The pooled odds ratios for citalopram, escitalopram and venlafaxine were not significant (figure 4). Removal of the outlying data in the citalopram analysis resulted in a statistically significant odds ratio of 1.28 (95% CI: 1.12 to 1.46, p = 0.0002), reported in supplement 5.

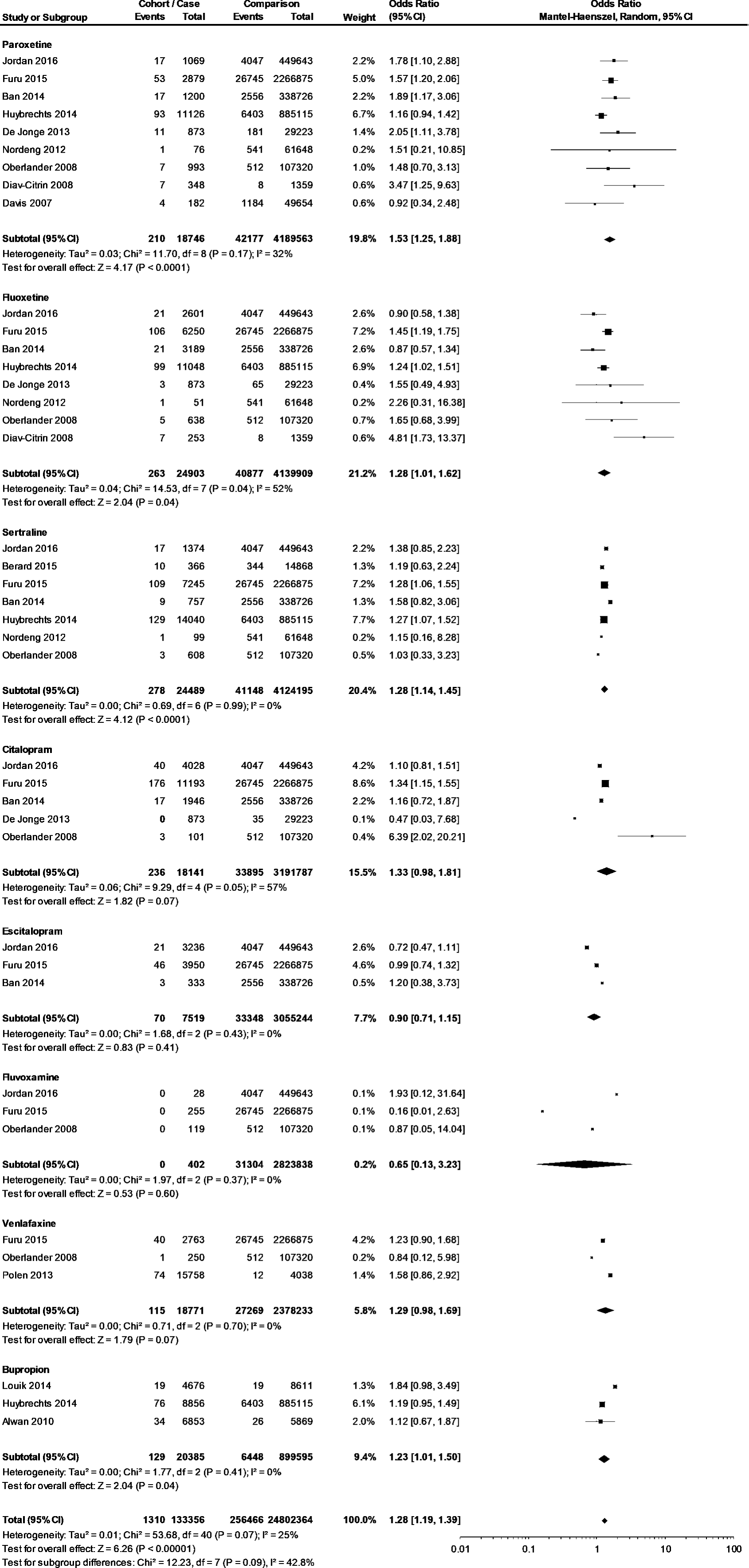

Analyses of cohort versus case-control studies resulted in odds ratios of 1.19 (95% CI: 1.08 to 1.31, p = 0.003) and 1.48 (95% CI: 1.04 to 2.11, p = 0.03), respectively (figure 5). Analyses of prospective versus retrospective studies resulted in odds ratios of 1.21 (1.09 to 1.34, p = 0.0005) and 1.24 (95% CI: 0.96 to 1.59, p = 0.10), respectively (figure 6).

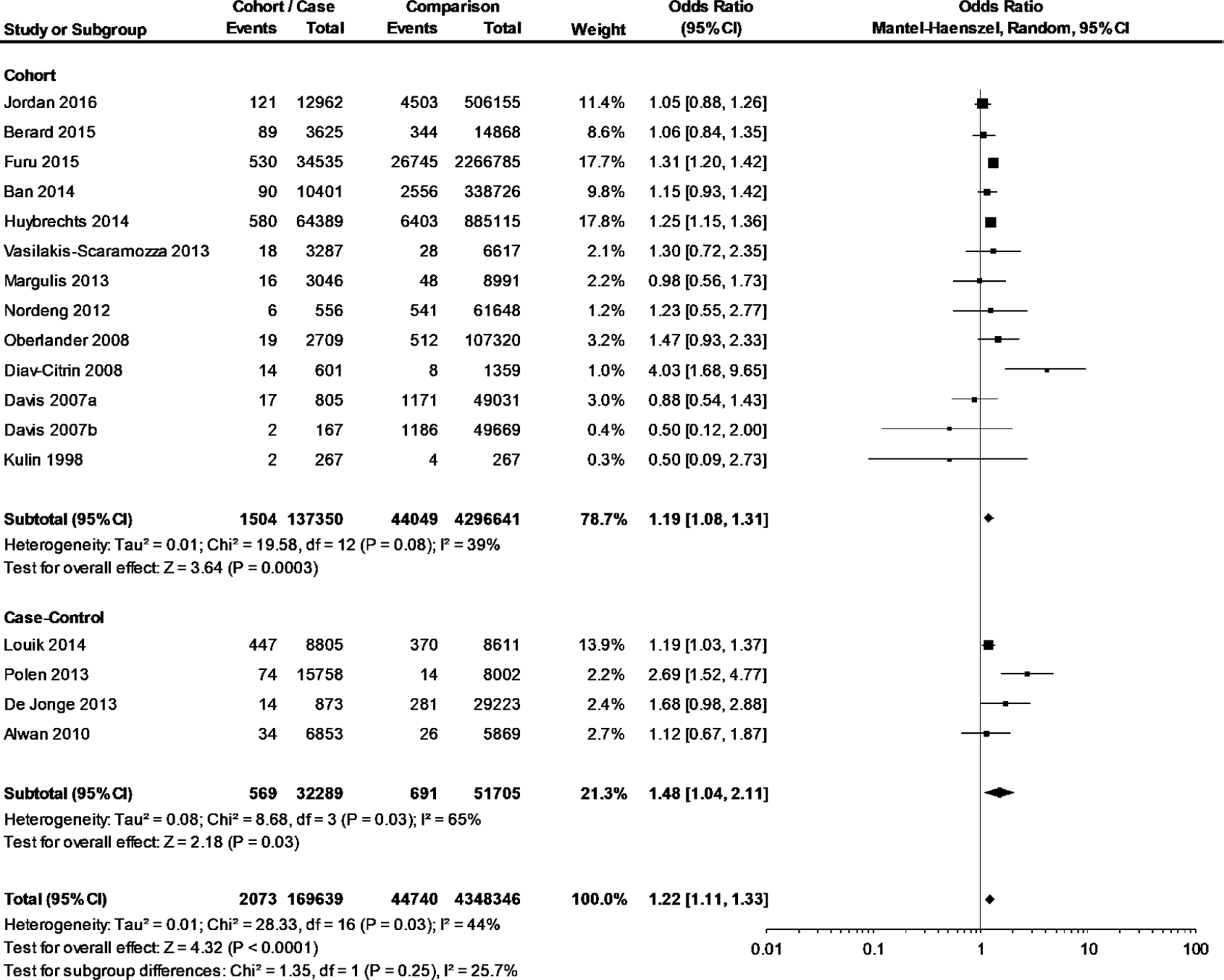

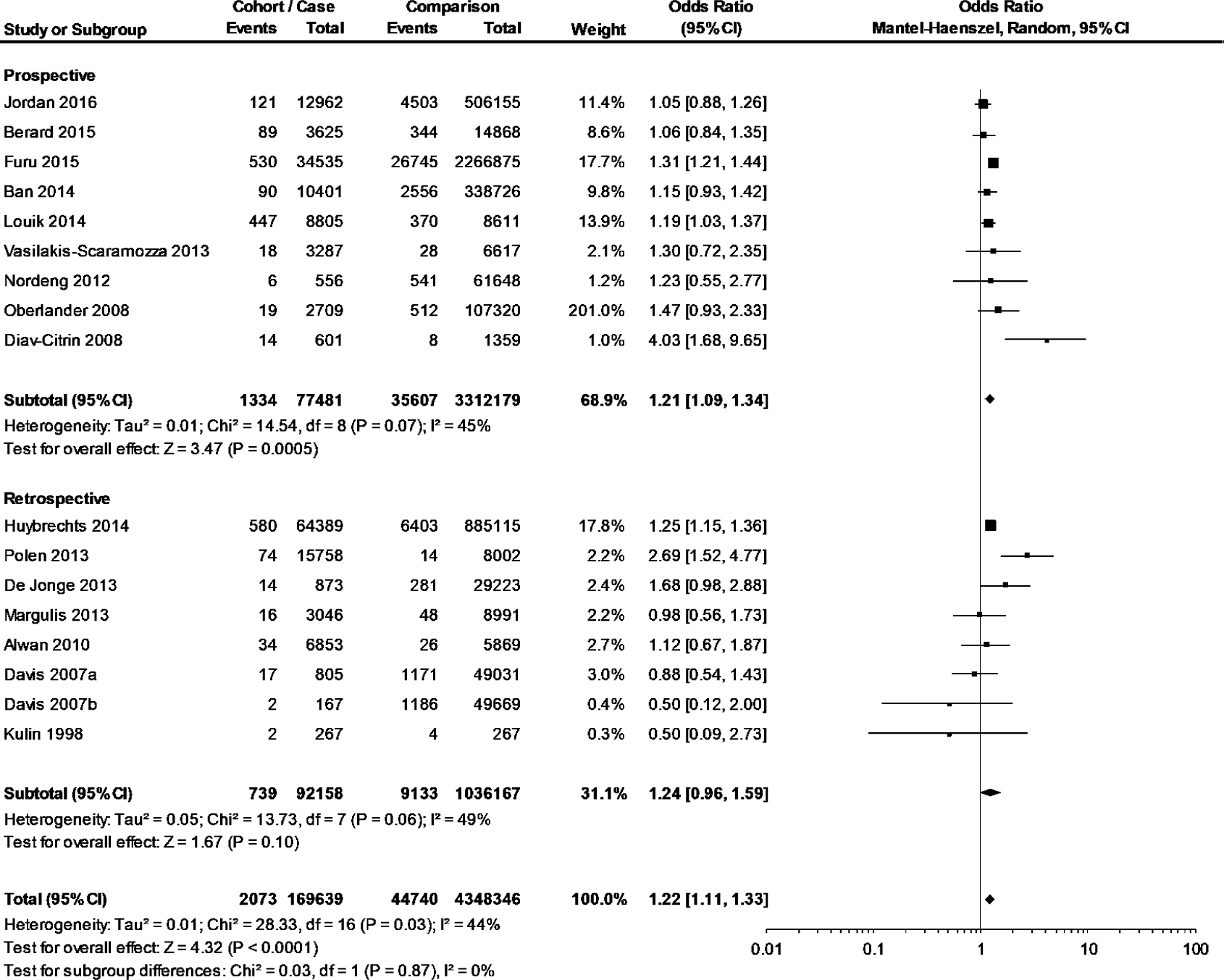

## Discussion

This meta-analysis of 16 studies from 13 countries including 4,564,798 pregnancy outcomes spanning twenty years confirms that using antidepressants during the first trimester of pregnancy is associated with an increased risk of congenital heart defects. Specifically, the odds of women who use any antidepressant in their first trimester of pregnancy having children with congenital heart defects is 1.1 to 1.3 times greater than the unmedicated group, at a 95% confidence level.

Our results for the usage of any antidepressant^13^, paroxetine,^5 8 9^ fluoxetine,^10,11^ and sertraline^12^ are comparable to previous meta-analyses, an overview of which is provided in table 3. These results indicate that both SNRIs and SSRIs increase the odds of congenital heart defects following maternal usage of these antidepressants during the first trimester of pregnancy, the critical period for cardiac development. Additionally, no increased odds for congenital heart defects following the usage of TCAs during the first trimester of pregnancy were found. This is the first meta-analysis to investigate and compare non-SSRI classes of antidepressants. These data are useful for providing insight into class-effects and possible mechanistic similarities for the formation of heart defects, as both SNRIs and SSRIs inhibit serotonin reuptake.^34,35^ However, limitations still exist in that there is still a lack of data about commonly used antidepressants, as demonstrated from Australian data; statistics for dispensed medications in 2013 identified that 16% of women of child bearing age who were dispensed psycho-analeptics were dispensed the SNRI desvenlafaxine. While our study investigated maternal SNRI usage during pregnancy and congenital heart defects, there was limited data on specific SNRIs and none of the studies investigating SNRIs had specific data on desvenlafaxine.^16 23 24 28 32^

**Table 3.** Previous meta-analyses assessing the relationship between antidepressant usage during the first trimester of pregnancy and cardiovascular defects

| **Author, Year Published** | **Data Sources (Search Period)** | **Study Types (Setting)** | **Tool For Assessing Study Quality** | **N Pregnant Women** | **Antidepressants Investigated** | **Reported OR/RR (95% CI)** | **I<sup>2</sup> (p Value)** | **Study Strengths** | **Study Weaknesses** |
| --- | --- | --- | --- | --- | --- | --- | --- | --- | --- |
| Shen, et al. 2017<sup>12</sup> | PubMed and Web of Science (inception to 31 December 2015) | 12 cohort studies (Europe, Australia, North and South America, Israel) | Newcastle Ottawa Scale | 6,468,241 | Sertraline | 1.36 (1.06 to 1.74) | 64.4% (0.001) | Limited to cohort studies<br>Modest power size and population heterogeneity explored<br>One antidepressant only | Only live births considered<br>Case-control studies not included<br>Moderate heterogeneity<br>Overlapping study data<br>Comparison group could have used other antidepressants |
| Gao, et al. 2017<sup>10</sup> | PubMed and Web of Science (inception to 21 March 2016) | 12 cohort studies (Europe, North America, Australia, Israel) | Newcastle Ottawa Scale | 6,450,771 | Fluoxetine | 1.36 (1.17 to 1.59) | 23.4% (0.214) | Limited to cohort studies and population heterogeneity explored<br>Multiple defects investigated<br>One antidepressant only<br>Live births, fetal deaths and terminations of pregnancy included.<br>Low heterogeneity observed in results | Case-control studies not included |
| Berard, et al. 2016<sup>5</sup> | Embase and Medline (1996 to November 2015) | 10 studies, cohort and case-control (North America, Europe, Australia) | Inclusion criteria: paroxetine use during T1, a comparator group, information to calculate an OR or a given RR or OR, investigated | 24,303 women exposed to paroxetine | Paroxetine | 1.23 (1.06 to 1.43) | 1.0% (0.436) | Limited to case-control and cohort studies<br>One antidepressant only<br>Low heterogeneity observed in results<br>Live births, fetal deaths and terminations of pregnancy included. | Comparison group could have used other antidepressants |

| congenital malformations |  |  |  |  |  |  |  |  |  |
| --- | --- | --- | --- | --- | --- | --- | --- | --- | --- |
| Riggin, et al. 2013<sup>11</sup> | PubMed, Medline and Embase (inception to 31 August 2012) | 15 cohort studies and 2 case-control studies | Inclusion criteria: any fluoxetine use during T1, compared to a control group not using SSRIs | Cohort: 3,437,257<br>Case-control: 23,452 | Fluoxetine | Cohort: 1.60 (1.31 – 1.95)<br>Case-control: 0.63 (0.39 – 1.03) | n.d. (0.431)<br>n.d. (0.14) | Limited to case-control and cohort studies<br>One antidepressant only<br>Three databases investigated<br>Include data outside of live births | Unclear on study procedures<br>Some control groups may have been exposed to non-SSRI antidepressants |
| Myles, et al. 2013<sup>8</sup> | CINAHL, Embase, Medline, PsycINFO and ISI Web of Science (1985 to June 2011) | 15 cohort studies | Based on Meta-Analysis Of Observational Studies in Epidemiology | 2,426,925 | Fluoxetine | 1.25 (0.98 – 1.60) | 33% (0.19) | Individual antidepressants investigated<br>Population heterogeneity explored<br>5 databases investigated<br>Includes data outside of live births | Limited to SSRIs only |
|  |  |  |  |  | Paroxetine | 1.44 (1.12 – 1.86) | 0% (0.82) |  |  |
|  |  |  |  |  | Sertraline | 0.93 (0.70 – 1.24) | 63% (0.03) |  |  |
|  |  |  |  |  | Citalopram | 1.03 (0.80 – 1.32) | 0% (0.58) |  |  |
| Grigoriadis, et al. 2013<sup>13</sup> | Embase, CINAHL, PsycINFO and Medline (inception to June 2010) | 13 studies, cohort and case-control | Systematic Assessment of Quality in Observational Research | 1,547,012 | Any antidepressant | 1.36 (1.08 – 1.71) | 31.1% (0.134) | Live births, fetal deaths and terminations of pregnancy included | Considers multiple antidepressants<br>Four of 13 studies had comparison groups which had used antidepressants |
| Bar-Oz, et al. 2007<sup>9</sup> | Medline, Embase, Reprotox, Scopus and Biological Abstracts (1985 to 2006) | 6 studies: 1 nested case control, 2 prospective controlled, 1 population-based cohort, 1 retrospective cohort, 1 prospective recording registry | Downs and Black | 16,789 | Paroxetine | 1.72 (1.22 – 2.42) | n.d. | 5 data sources investigated | Small number of pregnant women<br>Includes abstracts and a letter<br>3 control groups used other SSRIs<br>Heterogeneity not investigated<br>Live births only |

OR, odds ratio; RR, relative risk; CI, confidence interval; T1, First trimester of pregnancy; SSRI, selective serotonin reuptake inhibitor; n.d., no data

Venlafaxine and bupropion were the only SNRIs for which there was enough data to include in the individual antidepressant meta-analysis (figure 4), with respective pooled odds ratios of 1.29 (95% CI: 0.98 to 1.69) and 1.23 (95% CI: 1.01 to 1.50). Bupropion is not registered as an antidepressant in

Australia and is instead used for nicotine dependence and the dose size for this is 150 mg. Comparatively, the reported mean dose of bupropion in the USA from the Louik et al.^18^ study ranged from 254mg in monotherapy to 354mg in polytherapy.

SSRIs have been the most investigated class of antidepressants in relation to congenital heart defects, probably as a result of the 2005 paroxetine warning.^4^ Significant odds ratios for paroxetine, fluoxetine and sertraline were found (figure 4). Previous meta-analyses have documented a statistically significant relationship between maternal use of paroxetine,^5 8 9^ fluoxetine,^10 11^ and sertraline^12^ during the first trimester of pregnancy and congenital heart defects.

The pooled odds ratios for citalopram and escitalopram were not significant (figure 4) and a 2013 meta-analysis also calculated a non-significant pooled odds ratio for maternal citalopram usage and congenital heart defects.^8^ The removal of outlying data in the citalopram analysis resulted in a statistically significant odds ratio of 1.22 (1.03 to 1.45), therefore it is possible that citalopram may increase the likelihood for congenital heart defects (supplement 5); further escitalopram had a limited sample size.

The largest previous meta-analysis was carried out by Shen et al.^12^ and investigated the first trimester use of sertraline and congenital heart defects. Their meta-analysis included 12 cohort studies and had a total of 6,468,241 pregnant women, which is larger than the 4,564,798 pregnancy outcomes in this meta-analysis. We used six of the same studies^14-16 23 27 28^ however the remaining six from the Shen et al. meta-analysis were excluded for the following reasons: the Colvin et al. 2011^37^ study had a comparison group which had potentially used non-SSRI antidepressants; the Merlob et al. 2009^38^ study did not meet our inclusion criteria primarily because of self-reported SSRI usage and an unblinded assessment of pregnancy outcomes; the remaining four studies^39-42^ all reported data from the Swedish birth registry and these data therefore overlap with the Furu 2015 study (supplement 4).^16^ Like the Shen et al. ^12^ meta-analysis, our results for sertraline use during the first trimester of pregnancy and congenital heart defects are significant, while demonstrating lower heterogeneity, reflected by the I^2^ value (figure 4, table 3).

The Davis et al. 2007^21^ tricyclic antidepressant data and the Kulin et al. 1998^25^ study were classified as outliers due to their wide variances. The Diav-Citrin et al. 2008^33^ and Polen et al. 2013^32^ studies were classified as outliers because they demonstrated markedly higher intervention effect estimates than the overall odds ratio. Removal of these studies did not change the significance of the results but did decrease the heterogeneity (supplement 5).

Results were stratified by study type because it has been established that cohort studies are more rigorous than case-control studies as the study design results in less potential for sampling, observation and recall bias.^43 44^ Both cohort and case-control studies may be affected by confounding factors, including maternal usage of other medications and maternal smoking. The pooled average odds ratios were similar between both study groups (figure 5). Similarly, prospective studies are more rigorous than retrospective studies because these are conducted before pregnancy outcomes are known, resulting in more control over study features and less potential for recall bias.^43 44^ The pooled average odds ratios were significant for the prospective studies and not significant for the retrospective studies (figure 6).

A strength of our study was the inclusion criteria requiring all studies to be of high quality. Therefore, the meta-analyses reported a direct comparison between women who did or did not use antidepressants during the first trimester of pregnancy. One difficulty in generating high quality pharmacoepidemiological data is proper documentation of data in medical records and administrative databases, because there may be missing data, misclassification of medication usage or birth outcomes. One limitation of this meta-analysis is confirming whether the women used the antidepressants. Some studies confirmed this by structured interviews,^18 25 27 30 33^ while others only relied on antidepressant dispensings which do not verify consumption.^16 21 23 24 28 29^

A strength of this meta-analysis was that it was not limited to studies which only investigate live births. Live birth studies are susceptible to selection bias because congenital heart defects can be detected by scans which generally occur between weeks 18 to 22 of pregnancy.^45^ Pregnancies may be terminated if these defects are detected and these cases or events will be missed if only live births are assessed. Additionally, congenital heart defects may be present in stillbirths. Therefore, restricting analyses to live births may underestimate the relationship between maternal antidepressant use and congenital heart defects. Despite the potential for selection bias, two of the previous meta-analyses restricted pregnancy endpoints to live births only. ^9 12^ Similar to the results reported here, these meta-analyses found significant results for paroxetine^9^ and sertraline^12^ usage and heart defects, however their odds ratios were slightly larger and have wider confidence intervals (figure 4, table 3).

Our study, limited to high-quality studies, updates the literature on all antidepressants, including antidepressants belonging to the SNRI and TCA classes. There is evidence that maternal use of SNRIs during the first trimester of pregnancy increases the likelihood for congenital heart defects. Additionally, this meta-analysis confirms evidence for the SSRI class having an increased likelihood for causing cardiac defects. Some individual SSRIs, paroxetine, fluoxetine and sertraline, demonstrated statistically significant odds ratios. A statistically significant odds ratio was also demonstrated for the SNRI bupropion. No statistically significant odds ratio was produced for maternal usage of TCAs in the first trimester of pregnancy and congenital heart defects. While this meta-analysis has investigated classes of antidepressants and individual antidepressants in the context of congenital heart defects, further information is needed for some individual antidepressants which are used by women of child-bearing age, such as desvenlafaxine.

## Meanings Of Study

This meta-analysis has demonstrated that using antidepressants during the first trimester of pregnancy increases the overall risk for congenital heart defects. TCAs were the only group of antidepressants investigated in this study which did not show an increased odds ratio between maternal use and congenital heart defects. In comparison, SNRIs and SSRIs demonstrated an approximate increased likelihood for congenital heart defects of 1.5 and 1.2 times that of the unmedicated comparison group, respectively.

Analyses of individual antidepressants revealed that the SSRIs paroxetine, fluoxetine and sertraline increased the likelihood of congenital heart defects by 1.5, 1.3 and 1.3 times that of the comparison group respectively. Analyses of individual antidepressants revealed that the SNRI bupropion increased the likelihood of congenital heart defects by 1.2 times that of the comparison group.

Less information is available for some antidepressants such as desvenlafaxine of fluvoxamine, despite desvenlafaxine being the most dispensed psycho-analeptic medication in women of child-bearing age in Australia in 2013.^36^ It is also necessary to investigate risks associated for fluvoxamine because even though it is not as commonly used, medications may become more popular over time or be repurposed.

This meta-analysis has investigated the likelihood of antidepressants causing congenital heart defects however these medications may have other adverse effects which should also be investigated. Therefore, care should be taken when prescribing antidepressant medications to women who are likely to become pregnant, as switching or withdrawing antidepressant therapy is a slow process. Antidepressant therapy can also be ceased prior to conception for women who are well and perhaps more of an emphasis should be placed upon withdrawing medications if they are not needed.

## Future Directions

More pharmacoepidemiological data is needed to build a case for the risk of individual antidepressants causing congenital heart defects, especially those which have not been well studied such as desvenlafaxine and fluvoxamine. Ideally, analyses of all individual antidepressant medications should be completed in the context of other harmful pregnancy outcomes to identify which antidepressants present less risks during pregnancy than others.

If biological targets which play a key role in cardiac formation could be identified, this information could be used to detect medications which may cause congenital heart defects. Firstly, genes involved in cardiac defects can be identified from online sources, such as the Comparative Toxicogenomics Database (http://ctdbase.org/). The most promising genes can be investigated to determine if medications can directly interact with them and molecular modelling may be undertaken, particularly if X-ray crystal structures exist. Medications of interest can be computationally docked into the appropriate binding site to enable potential identification of antidepressants or other medications which may cause congenital heart defects. Laboratory luciferase assays may provide additional evidence for any suspected harmful medications which are identified.

The aim of future work is therefore to identify which medicines may cause congenital heart defects by investigating the mechanisms behind medication-related cardiac malformations.

## Conclusions

Maternal usage of antidepressants during the first trimester of pregnancy increases the risk for congenital heart defects by approximately 1.2 times that of the group not using antidepressants during pregnancy. SNRI and SSRI antidepressants showed an increased risk of 1.5 and 1.2 times respectively, while TCAs were not found to have an increased risk. Individual SSRIs paroxetine, fluoxetine and sertraline have demonstrated an approximate increased likelihood of congenital heart defects by 1.5, 1.3 and 1.3 times respectively. The SNRI bupropion also demonstrated an approximate 1.2 times increased likelihood of congenital heart defects.

More information is needed about the risks of individual antidepressants, especially for desvenlafaxine, the most dispensed psycho-analeptic in Australia during 2013. The mechanism of antidepressant-related heart defects is currently unknown and future computational and laboratory work could be undertaken to understand these mechanisms and to potentially identify other medications which increase the risk of congenital heart defects.

## Data Availability

Additional data is provided in the supplementary files.

## Footnotes

### Contributors

ER and CDV designed the study. CDV did the literature search. CDV and SG extracted data from the identified articles. CDV did the meta-analyses and drafted the manuscript. All authors reviewed the manuscript, contributed to its revision and approved the final version submitted.

### Funding

This work was supported by an Australian Government Research Training Program scholarship and NHMRC APP 1110139.

### Competing interests

All authors have completed the ICMJE uniform disclosure form at and declare: no support from any organisation for the submitted work; the University of South Australia received payments from the Australian Government Research Training Program, which is a PhD student scholarship, with no financial relationships with any other organisations that might have an interest in the submitted work in the previous three years, excepting no other relationships or activities that could appear to have influenced the submitted work.

### License statement

CDV has the right to grant on behalf of all authors and does grant on behalf of all authors, a worldwide licence to the Publishers and its licensees in perpetuity, in all forms, formats and media (whether known now or created in the future), to i) publish, reproduce, distribute, display and store the Contribution, ii) translate the Contribution into other languages, create adaptations, reprints, include within collections and create summaries, extracts and/or, abstracts of the Contribution and convert or allow conversion into any format including without limitation audio, iii) create any other derivative work(s) based in whole or part on the on the Contribution, iv) to exploit all subsidiary rights to exploit all subsidiary rights that currently exist or as may exist in the future in the Contribution, v) the inclusion of electronic links from the Contribution to third party material where-ever it may be located; and, vi) licence any third party to do any or all of the above. All research articles will be made available on an open access basis (with authors being asked to pay an open access fee). The terms of such open access shall be governed by a Creative Commons licence—details as to which Creative Commons licence will apply to the research article are set out in our worldwide licence referred to above.

### Ethical approval

Not needed.

### Data sharing

Additional data is provided in the supplementary files.

### Transparency

The corresponding author affirms that the manuscript is an honest, accurate, and transparent account of the study being reported; that no important aspects of the study have been omitted; and that any discrepancies from the study as planned have been explained.

### Patient and Public Involvement Statement

Patients or the public were not involved in the design, or conduct, or reporting, or dissemination plans of our research.

### Dissemination Declaration

Dissemination to study participants or patient organisations is not possible/applicable.

